# Newborn Antibodies to SARS-CoV-2 detected in cord blood after maternal vaccination

**DOI:** 10.1101/2021.02.03.21250579

**Authors:** Paul Gilbert, Chad Rudnick

## Abstract

**Background:** Maternal vaccination for Influenza and TDaP have been well studied in terms of safety and efficacy for protection of the newborn by placental passage of antibodies. Similar newborn protection would be expected after maternal vaccination against SARS-CoV-2 (the virus responsible for COVID-19). There is a significant and urgent need for research regarding safety and efficacy of vaccination against SARS-CoV-2 during pregnancy. Here, we report the first known case of an infant with SARS-CoV-2 IgG antibodies detectable in cord blood after maternal vaccination.

**Case presentation:** A vigorous, healthy, full-term female was born to a COVID-19 naïve mother who had received a single dose of mRNA vaccine for SARS-CoV-2 three weeks prior to delivery. Cord blood antibodies (IgG) were detected to the S-protein of SARS-CoV-2 at time of delivery.

**Conclusion:** Here, we report the first known case of an infant with SARS-CoV-2 IgG antibodies detectable in cord blood after maternal vaccination.

## Background

Maternal vaccination for Influenza and TDaP have been well studied in terms of safety and efficacy for protection of the newborn by placental passage of antibodies. Similar newborn protection would be expected after maternal vaccination against SARS-CoV-2 (the virus responsible for COVID-19). There is a significant and urgent need for research regarding safety and efficacy of vaccination against SARS-CoV-2 during pregnancy. Here, we report the first known case of an infant with SARS-CoV-2 IgG antibodies detectable in cord blood after maternal vaccination.

## Methods

Maternal vaccination was provided to a COVID-19-naïve front-line healthcare worker with the Moderna mRNA COVID-19 vaccine, at gestational age of 36 weeks 3 days. A normal, spontaneous vaginal birth occurred 3 weeks after dose 1 of the Moderna vaccine. The product of this 39 week 3 days gestation was a vigorous, healthy, full-term girl with normal newborn nursery course and subsequent well-infant evaluation. Under aseptic conditions and along with standard cord blood sampling for newborn blood type and direct antiglobulin test (DAT) a cord blood sample was taken immediately after birth and prior to placenta delivery with 0.5mL drawn into a red-top tube for serum. The serum was sent for SARS-CoV-2 antibody test to the S-protein as performed by Labcorp. The mother, who has been breastfeeding exclusively, then received the second dose of the Moderna vaccine during the post-partum period per the normal 28-day vaccination protocol timeline.

## Results

Cord blood antibodies (IgG) were detected to SARS-CoV-2 at a level of 1.31 U/mL. This Electrochemiluminescence Immunoassay (ECLIA) uses a recombinant protein representing the RBD (receptor-binding domain) of the S antigen for the quantitative determination of antibodies against SARS-CoV-2^1^.

## Discussion

Vaccination during pregnancy with TDaP and Flu is both well studied and formally recommended^2-5^. The COVID-19 Pandemic and subsequent EUAs for two mRNA vaccines have created significant need for active research regarding safety and efficacy in pregnant and lactating mothers, as well as their offspring^6,7^.

The novel mRNA vaccines theoretically will demonstrate similar safety in this population, including placental passage of protective antibodies. Natural SARS-CoV-2 infection, however, seems to confer lower than expected passage of antibodies to the fetus which may indicate newborns born to vaccinated mothers will remain at risk for infection^8^.

We have demonstrated that SARS-CoV-2 IgG antibodies are detectable in a newborn’s cord blood sample after only a single dose of the Moderna COVID-19 vaccine. Thus, there is potential for protection and infection risk reduction from SARS-CoV-2 with maternal vaccination. Quantification of the antibody response can help to determine the specific antibody titer and aid in longitudinal monitoring of the dynamics of the antibody response in individual patients. Protective efficacy in newborns and ideal timing of maternal vaccination remains unknown. Further studies will be needed to quantify the amount of viral neutralizing antibodies present in babies born to SARS-CoV-2 naïve mothers who are vaccinated prior to delivery. Additionally, the duration of antibody protection in this population is not yet known and serial total antibody measurements may be used to determine how long protection is expected which may help to determine when the best time would be to begin vaccination in newborns born to mothers who received a vaccine for SARS-CoV-2.

We urge other investigators to create pregnancy and breastfeeding registries as well as conduct efficacy and safety studies of the COVID-19 vaccines in pregnant and breastfeeding woman and their offspring.

## Data Availability

data referred to in the manuscript is available at request.

